# Cohort profile: The Swedish Inception Cohort in inflammatory bowel disease (SIC-IBD)

**DOI:** 10.1101/2025.01.12.25320315

**Authors:** Benita Salomon, Olle Grännö, Daniel Bergemalm, Hans Strid, Adam Carstens, Henrik Hjortswang, Maria Ling Lundström, Jóhann P Hreinsson, Sven Almer, Francesca Bresso, Carl Eriksson, Olof Grip, André Blomberg, Jan Marsal, Niloofar Nikaein, Shoaib Bakhtyar, Carl Mårten Lindqvist, Elisabet Hultgren Hörnquist, Maria K Magnusson, Åsa V Keita, Mauro D’Amato, Dirk Repsilber, Lena Öhman, Johan D Söderholm, Marie Carlson, Charlotte R H Hedin, Robert Kruse, Jonas Halfvarson

## Abstract

**Purpose:** There is a need for diagnostic and prognostic biosignatures to improve long-term outcomes in inflammatory bowel disease (IBD). Here, we describe the establishment of the Swedish inception cohort in IBD (SIC-IBD) and demonstrate its potential for the identification of such signatures.

**Participants:** Patients aged ≥18 years with gastrointestinal symptoms who were referred to the gastroenterology unit due to suspected IBD at eight Swedish hospitals between November 2011 and March 2021 were eligible for inclusion.

**Findings to date:** In total, 368 patients with IBD (Crohn’s disease, n=143; ulcerative colitis, n=201; IBD-unclassified, n=24) and 168 symptomatic controls were included. In addition, 59 healthy controls without gastrointestinal symptoms were recruited as a second control group. Biospecimens and clinical data were collected at inclusion and in patients with IBD also during follow-up to 10 years. Levels of faecal calprotectin and high-sensitivity C-reactive protein were higher in patients with IBD compared to symptomatic controls and healthy controls. Preliminary results highlight the potential of serum protein signatures and autoantibodies, as well as results from faecal markers, to differentiate between IBD and symptomatic controls in the cohort. During the first year of follow-up, 36% (52/143) of the patients with Crohn’s disease, 24% (49/201) with ulcerative colitis and 4% (1/24) with IBD-U experienced an aggressive disease course.

**Future plans:** We have established an inception cohort enabling ongoing initiatives to collect and generate clinical data and multi-omics datasets. The cohort will allow analyses for translation into candidate biosignatures to support clinical decision-making in IBD. Additionally, the data will provide insights into mechanisms of disease pathogenesis.

**Bullet points on strengths and weaknesses:**

- This large, multicentre inception cohort of newly diagnosed adult IBD patients, with integrated biobanking and prospective material collection, enables exploration of IBD pathophysiology and biomarker discovery.
- The inclusion of symptomatic controls with gastrointestinal symptoms mimicking IBD but without evidence of the diagnosis provides a realistic diagnostic setting and addresses limitations of many previous cohorts.
- The use of healthy controls as a second control group offers opportunities to gain insight into IBD pathogenesis.
- The non-population-based study design and the predominance of university hospitals in recruitment may limit the generalisability of findings.
- Suboptimal sample processing at some centres, including delays in serum centrifugation and transportation of faecal samples at ambient temperatures, may have affected the quality of some specific analyses, such as microbiota analyses.

## INTRODUCTION

Inflammatory bowel disease (IBD), comprising Crohn’s disease, ulcerative colitis and IBD-unclassified (IBD-U), is a chronic disease that predominantly presents during the second and third decades of life. The disease results in an impaired quality of life for patients and substantial societal costs due to sick leave and a high use of healthcare resources [1–3]. Progressive inflammation causes damage to the gastrointestinal tract, and a substantial proportion of patients develop serious disease complications, including bowel obstruction, fistula, abscesses, and colorectal cancer [4–6].

IBD often presents with non-specific symptoms such as diarrhoea, fatigue and abdominal pain and, therefore, presents a diagnostic challenge. Commonly used biomarkers in diagnostic algorithms include C-reactive protein (CRP) and, in some regions, faecal calprotectin (FCP) which help identify patients needing referral for further investigations, including ileocolonoscopy [7]. However, the diagnostic accuracy of CRP is too low for reliably identifying IBD patients, and the utility of FCP is hampered by poor patient adherence to faecal sampling [8,9]. Furthermore, many healthcare systems do not reimburse the use of FCP for diagnostic purposes. The absence of reliable, non-invasive diagnostic biomarkers, occurrence of non-specific symptoms and limited access to ileocolonoscopy, frequently results in a significant diagnostic delay [10,11]. As a consequence, many patients have acquired considerable bowel damage (e.g., fistulas, strictures) already at diagnosis [4,11].

At diagnosis, the heterogeneity in clinical presentation and subsequent disease course make clinicians’ attempts to stratify the treatment according to the individual patient’s needs very difficult. While some patients experience an indolent disease course with minimal symptoms and low endoscopic activity, a substantial proportion exhibit an aggressive disease phenotype characterised by treatment refractoriness, corticosteroid dependency, frequent hospital admissions and surgical interventions, including resection surgery and acute colectomy [12–18]. For this latter group, an early effective treatment seems advantageous, as the initial disease phase may provide a “window of opportunity” where prompt suppression of intestinal inflammation could improve long-term outcomes [19–21]. Identifying prognostic biomarkers for this group of patients is therefore critical to facilitate the timely initiation of potent treatment.

To address the need for both diagnostic and prognostic biomarkers, we established the “Swedish Inception Cohort in IBD” (SIC-IBD), a large prospective multicentre cohort involving patients with suspected IBD referred to eight hospitals in Sweden. After a comprehensive diagnostic work-up, patients were classified as incident IBD patients or symptomatic non-IBD controls, i.e., patients presenting with symptoms mimicking IBD but where IBD was ruled out. Alternative diagnoses in this patient group included infectious colitis, irritable bowel syndrome and diverticulitis. As a second control group, healthy controls without gastrointestinal symptoms were recruited. This cohort provides a rich collection of biospecimens together with detailed phenotypic characterisation collected both at diagnosis and during follow-up. Herein, we present the study design and baseline characteristics of the participants in the SIC-IBD.

## COHORT DESCRIPTION

### Inclusion

This longitudinal multicentre cohort study was conducted across eight Swedish hospitals: Ersta Hospital, Karolinska University Hospital, Linköping University Hospital, Sahlgrenska University Hospital, Skåne University Hospital, Södra Älvsborg Hospital, Uppsala University Hospital, and Örebro University Hospital. Patients aged ≥18 years with gastrointestinal symptoms, such as diarrhoea, abdominal pain and blood or mucus in stools, who were referred to the gastroenterology unit with suspected IBD between November 2011 and March 2021 were eligible for inclusion (Figure 1). Exclusion criteria included a previous diagnosis of Crohn’s disease, ulcerative colitis or IBD unclassified, or inability to provide informed consent or to comply with protocol requirements. After obtaining written informed consent, all patients underwent a routine diagnostic work-up for IBD, following clinical practice and international guidelines [22].

**Figure 1.**
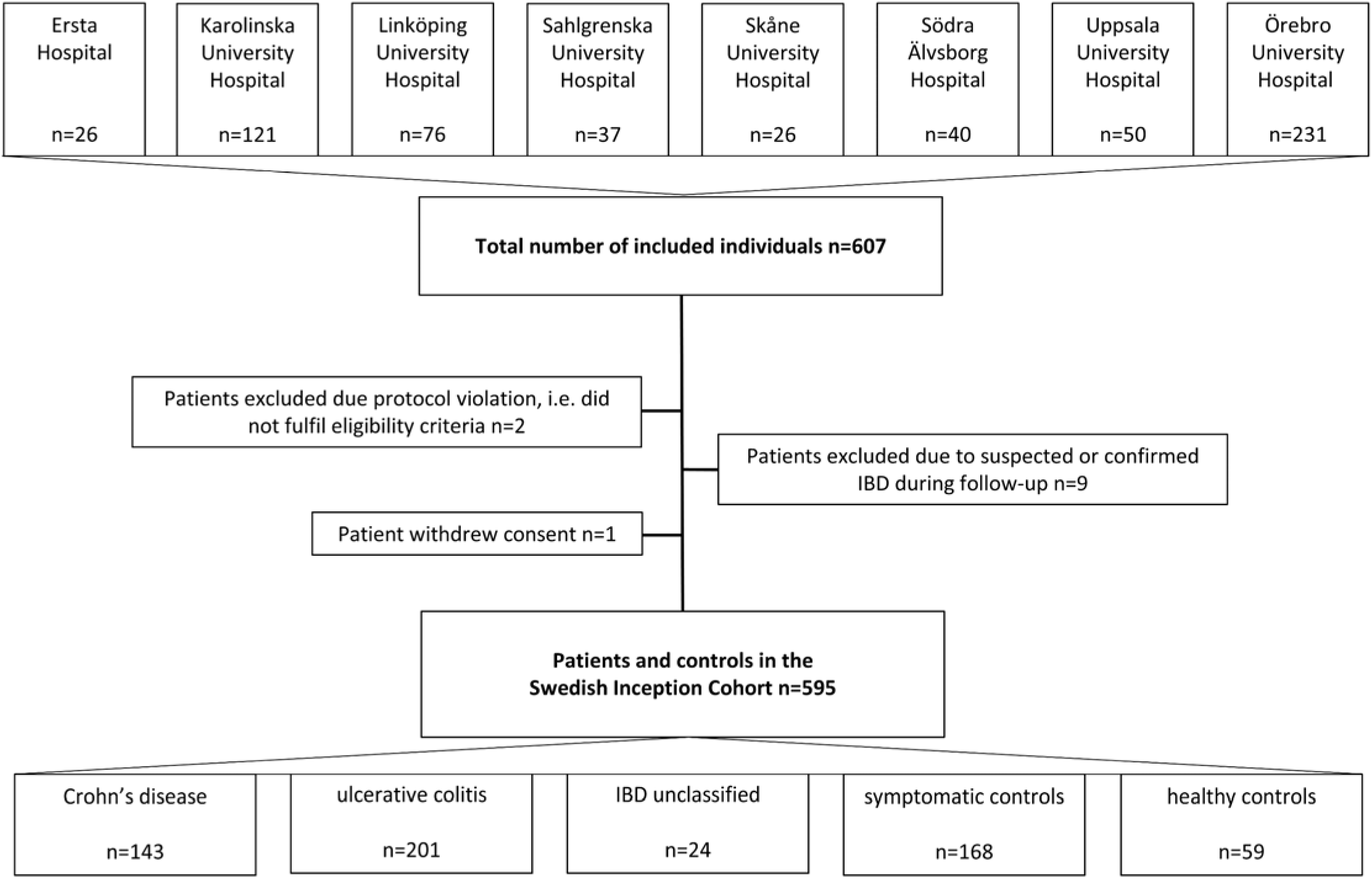
The flow diagram shows the number of included individuals at each hospital, the number of patients excluded and the final number of patients and controls in the Swedish Inception Cohort in inflammatory bowel disease (SIC-IBD).

Based on the diagnostic work-up, which included magnetic resonance imaging in cases where small bowel involvement was suspected, patients were classified as having incident IBD or as symptomatic controls, i.e., patients with no discernible evidence of IBD at inclusion or during follow-up. The diagnosis of IBD was based on internationally accepted clinical, endoscopic, radiological, and histological criteria [22]. Nine patients were excluded due to suspected or confirmed IBD during follow-up. Patients were followed prospectively according to clinical practice, with data collected after 3 months, 1, 3, 5, 7 and 10 years for those with IBD. In addition, a group of healthy controls, without gastrointestinal disease, was included at baseline.

### Demographics of patients and controls

During the inclusion period, 368 patients with IBD (Crohn’s disease, n=143; ulcerative colitis, n=201; IBD-U, n=24) and 168 symptomatic controls were included (**Table 1**). The group of symptomatic controls comprised patients who ultimately were diagnosed with various diseases, including microscopic colitis, infectious enteritis, celiac disease, or irritable bowel syndrome. In addition, 59 healthy controls without any history of gastrointestinal disease were recruited as a second control group.

**Table 1.** Demographics of individuals included in the Swedish Inception Cohort in IBD (SIC-IBD).

|  | **Crohn's**<br>**disease**<br>**n=143** | **Ulcerative**<br>**colitis**<br>**n=201** | **IBD-unclassified**<br>**n=24** | **Symptomatic**<br>**control**<br>**n=168** | **Healthy**<br>**control**<br>**n=59** |
| --- | --- | --- | --- | --- | --- |
| **Female sex, n (%)** | 73 (51.1) | 81 (40.3) | 11 (45.8) | 107 (63.7) | 33 (55.9) |
| **Median age (IQR)** | 28 (23-41) | 34 (26-45) | 33 (30-45) | 34 (25-48) | 26 (23-31) |
| **Median CRP, mg/L (IQR)\*** | 8.0 (3.3-31.8) | 3.6 (1.4-8.6) | 3.1 (2-5.6) | 1.8 (0.9-5.9) | 0.6 (0.3-1.1) |
| **Median FCP, µg/g (IQR)\*** | 448 (236-978) | 325 (90-880) | 471 (148-621) | 68 (19-146) | 10 (5-24) |
| **Smoking, n (%)** |  |  |  |  |  |
| Never | 70 (49) | 95 (48) | 12 (50) | 71 (42) | 41 (69) |
| Previous | 26 (18) | 59 (30) | 8 (33) | 29 (17) | 10 (17) |
| Current | 22 (15) | 15 (7) | 1 (4) | 16 (10) | 4 (7) |
| Missing | 25 (17) | 31 (16) | 3 (12) | 52 (31) | 4 (7) |
| **Median BMI, kg/m<sup>2</sup> (IQR)** | 23 (22-26) | 24 (22-27) | 27 (22-28) | 26 (22-30) | 23 (21-26) |

*\*Information was missing for CRP in 62 (10%) individuals, faecal calprotectin in 217 (37%)* *individuals and for Body Mass Index in 198 (33%) individuals*
*CRP: C-reactive protein; IQR; interquartile range; FCP, faecal calprotectin*

### Healthcare setting

Sweden has a tax-funded universal healthcare system, and private outpatient healthcare providers are few within the field of gastroenterology. In general, gastroenterologists at secondary or tertiary care centres (rather than primary care) are responsible for diagnosing and managing patients with IBD.

### Data collection

#### Characterisation of disease phenotype and disease activity

Following informed consent, data on patient demographics, disease phenotype according to the Montreal classification[23], extraintestinal manifestations, endoscopic activity (defined as the presence of ulcers in Crohn’s disease [yes/no] and the Mayo endoscopic subscore in ulcerative colitis), IBD treatment, IBD-related surgery, and hospital admission were recorded at baseline and prospectively during follow-up. Treatment-naivety was defined as no prior treatment with IBD-related medications, including topical therapies. Previous treatments in patients who were not treatment-naïve included mostly topical therapies but also a few days of corticosteroid treatment. For healthy controls, information about basic demographics was collected. Data on disease characteristics were recorded using case report forms (CRFs), and details on the Montreal classification and clinical disease activity at baseline are provided in **Table 2**.

**Table 2.** Clinical characteristics of patients with Crohn’s disease, ulcerative colitis and IBD-unclassified in the Swedish Inception Cohort in IBD (SIC-IBD).

|  |  | **Crohn's**<br>**Disease**<br>**n=143** | **Ulcerative**<br>**colitis**<br>**n=201** | **IBD-**<br>**unclassified**<br>**n=24** |
| --- | --- | --- | --- | --- |
| **Disease location, n (%)** |  |  |  |  |
|  | ileal (L1) | 48 (33) |  |  |
|  | colonic (L2) | 50 (35) |  |  |
|  | ileocolonic (L3) | 34 (24) |  |  |
|  | upper GI (L4) | 1 (1) |  |  |
|  | unknown | 10 (7) |  |  |
| **Disease behaviour, n (%)** |  |  |  |  |
|  | non-stricturing,<br>non-penetrating (B1) | 115 (80) |  |  |
|  | stricturing (B2) | 14 (10) |  |  |
|  | penetrating (B3) | 5 (3) |  |  |
|  | unknown | 9 (6) |  |  |
| **Perianal disease, n (%)** |  | 12 (8) |  |  |
| **Disease extent, n (%)** |  |  |  |  |
|  | ulcerative proctitis (E1) |  | 64 (32) |  |
|  | left-sided colitis (E2) |  | 57 (28) |  |
|  | extensive colitis (E3) |  | 76 (38) |  |
|  | unknown |  | 4 (2) |  |
| **Treatment-naïve, n (%)** |  | 128 (90) | 185 (92) | 21 (88) |

Upper GI (L4) refers to patients with only upper gastrointestinal Crohn's disease
IBD-unclassified, Inflammatory bowel disease unclassified; SIC-IBD; GI, gastrointestinal

#### Patient-reported outcomes

Patient questionnaires were employed to obtain information about clinical disease activity, smoking status, recent antibiotic or non-steroid anti-inflammatory drug use, and any specific dietary restrictions.

Patients were also asked to complete the Short-Health-Scale and Short Form-36 (SF-36) questionnaire to obtain information about Health-Related Quality of Life (HRQoL) [24–26]. SF-36 is a general measure of HRQoL, whereas the Short-Health Scale is a validated measure to address HRQoL in patients with IBD. It includes four dimensions: bowel symptoms, daily life activities, worry and general well-being. Additional quality of life data were collected using the EuroQol-5D (EQ-5D) questionnaire[27,28]. Mental health was evaluated using the Patient-health Questionnaire 9 (PHQ9) [29], the Cognitive Reserve Index (CRI) [30], and the Short-Form Health Survey (SF-36) [26]. Additional data on gastrointestinal symptoms was obtained by using the Gastrointestinal Symptom Rating Scale (GSRS) [31] and the Rome III criteria functional gastrointestinal disorders [32].

### Sample collection and biobanking

Biospecimens were obtained at baseline and during follow-up, processed and biobanked according to predefined standard operating procedures (SOPs). Blood samples were collected in EDTA and PAX®-tubes. In addition, serum samples were obtained by centrifugation of whole blood at 2400 × g for 5 min at ambient temperature, followed by aliquoting of serum and storing at −80° C. For six centres, blood samples were shipped unprocessed at ambient room temperature and centrifuged upon arrival at the central biobank, Örebro, Sweden. The participants collected faecal samples in plastic tubes at home, brought them to the hospital or mailed them by postal service. On arrival, the samples were immediately frozen and stored at −80°C. Spot urine samples were collected during study visits, aliquoted and stored at −80°C. In addition to collecting biopsies as part of clinical routine, physicians were instructed to take intestinal biopsies from both inflamed and non-inflamed mucosa according to a predefined protocol. These biopsies were preserved in Allprotect Tissue Reagent (Qiagen, Hilden, Germany) and in a bacterial freezing medium. Biopsies in the freezing medium were immediately stored at −80°C, whereas those in Allprotect Tissue Reagent were handled according to the manufacturer’s instructions. Details on the collected biological material are provided in **Table 3**.

**Table 3.**
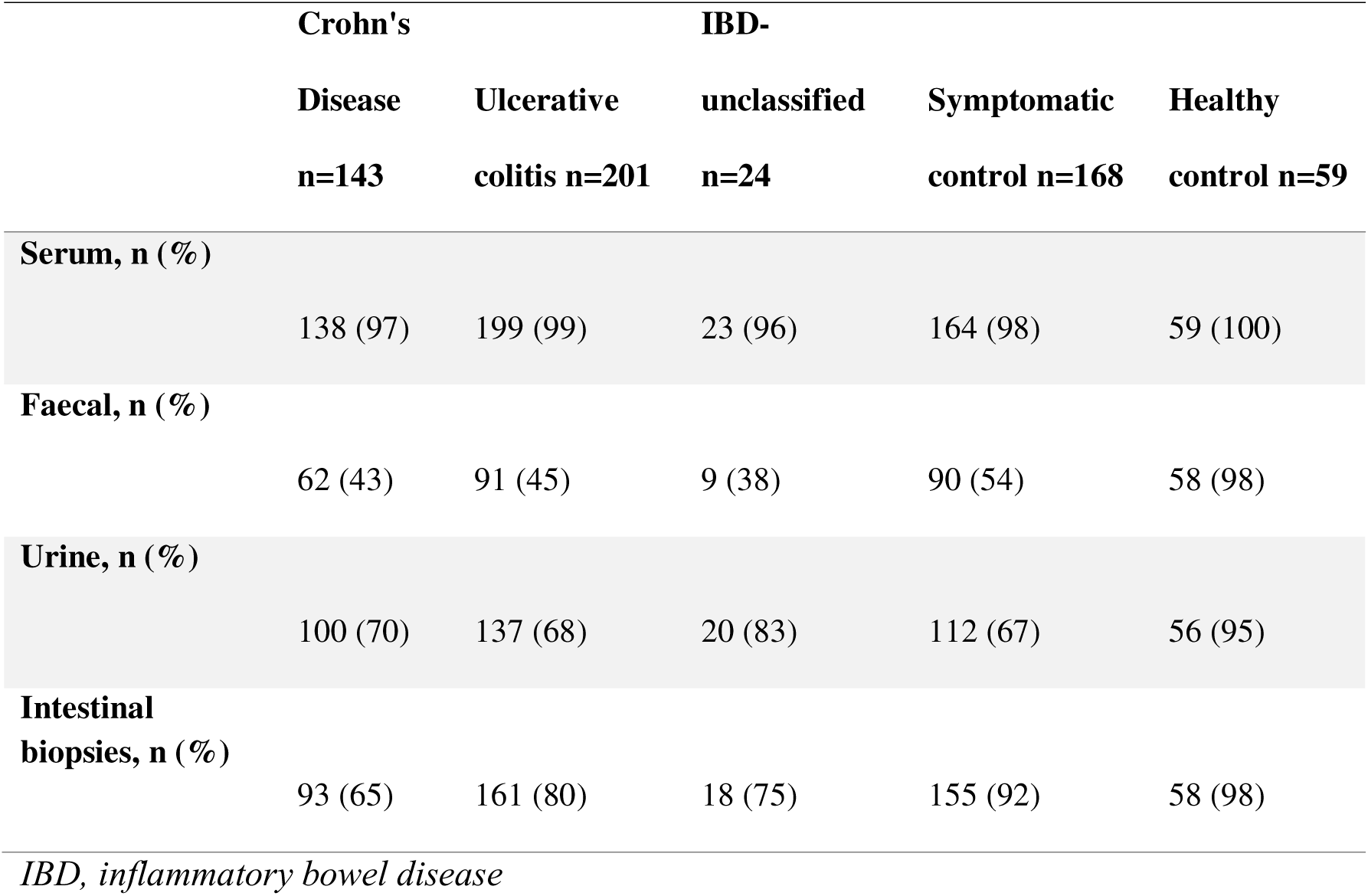
An overview of biospecimens collected at baseline from individuals within the Swedish Inception Cohort in inflammatory bowel disease (SIC-IBD).

|  | Crohn's |  | IBD- |  |  |
| --- | --- | --- | --- | --- | --- |
|  | Disease | Ulcerative | unclassified | Symptomatic | Healthy |
|  | n=143 | colitis n=201 | n=24 | control n=168 | control n=59 |
| **Serum, n (%)** |  |  |  |  |  |
|  | 138 (97) | 199 (99) | 23 (96) | 164 (98) | 59 (100) |
| **Faecal, n (%)** |  |  |  |  |  |
|  | 62 (43) | 91 (45) | 9 (38) | 90 (54) | 58 (98) |
| **Urine, n (%)** |  |  |  |  |  |
|  | 100 (70) | 137 (68) | 20 (83) | 112 (67) | 56 (95) |
| **Intestinal biopsies, n (%)** |  |  |  |  |  |
|  | 93 (65) | 161 (80) | 18 (75) | 155 (92) | 58 (98) |

*IBD, inflammatory bowel disease*

### Measurements of high-sensitivity CRP and faecal **c**alprotectin

After storage, samples for high-sensitivity (hs) CRP (serum) and calprotectin (faeces) were retrieved from the central biobank and sent to Uppsala Clinical Research Center, Uppsala, Sweden and Academic Laboratory, Department of Clinical Chemistry, University Hospital, Uppsala, Sweden, respectively. hsCRP was assayed in a single batch for all participants at the end of the recruitment period, using a particle-enhanced immunoturbidimetric hsCRP assay (Cardiac C-Reactive Protein (Latex) High Sensitive, Roche Diagnostics) on a Roche Cobas c501. Correspondingly, faecal calprotectin was extracted and analysed according to the manufacturer’s instructions, with a chemiluminescent immunoassay, using the LIASON XL analyser (DiaSorin, Saluggia, Italy). Data on median levels of hsCRP and FCP across the various diagnoses are provided in **Table 1**.

### Disease course outcome during follow-up

A composite outcome measure was used to categorise the disease course as aggressive or indolent during follow-up. An aggressive disease course was defined as the presence of any IBD-related surgery, hospital admission for active disease, treatment refractoriness towards targeted therapies, i.e., biologics, Janus kinase (JAK) inhibitors or sphingosine-1-phosphate receptor modulators, and >two courses or high cumulative dosis of systemic corticosteroids within the first year of follow-up (**Table 4**). In addition, Crohn’s disease patients with evidence of progression to a complicated disease behaviour, i.e., a new stricture, fistula, or abscess, were also categorised as having an aggressive disease course.

**Table 4.**
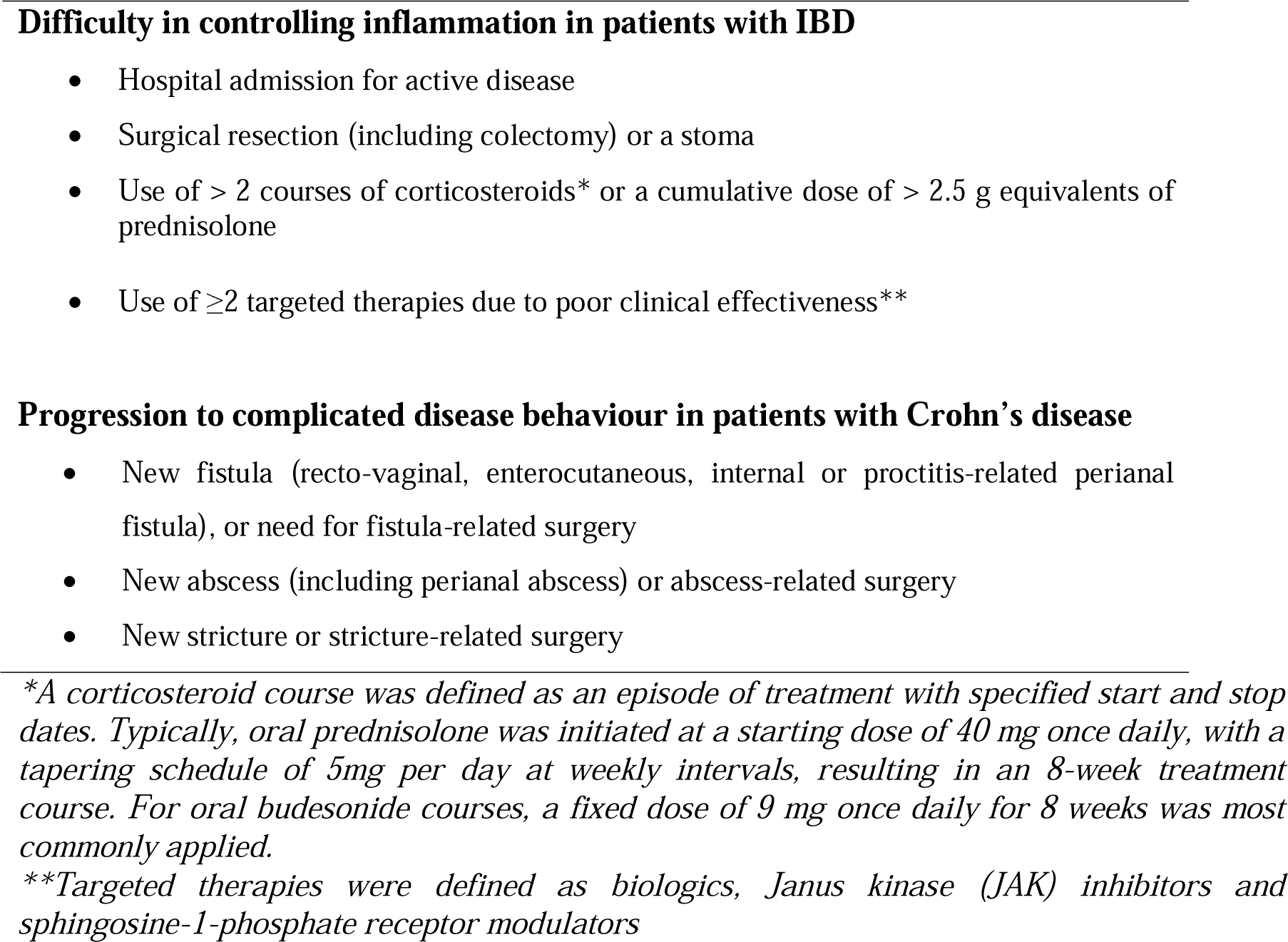
Criteria for defining the disease course as aggressive or indolent within the Swedish Inception Cohort in inflammatory bowel disease (SIC-IBD). Patients with any of the following events during the first year from the date of diagnosis were defined as having an aggressive disease course.

### Data management

Patients, physicians, or research nurses used CRFs and predefined questionnaires to collect information. All data were imported after pseudonymisation into a central database with reading access for all co-investigators. The database is updated every 6-12 months to allow the import of new data, including follow-up visits, address possible missing information, and correct potential erroneous data entries. To enable backward traceability, a data freeze is created at each update. Each data freeze provides a snapshot of information contained in the database at a given date.

### Project organisation

The SIC-IBD represents a crucial element of the multi-modal national study to identify biomarkers for diagnosis, therapy response and disease progression in IBD (BIO IBD), which aims to bring leading clinicians, clinical researchers and basic scientists within the field of IBD together with corporate partners. The managerial structure and the operational organisation of BIO IBD are shown in **Figure 2**. An executive office has been set up at the Faculty of Health and Medical Sciences, Örebro University, forming the BIO IBD Executive Committee together with representatives of participating universities and university hospitals. The Executive Committee coordinated the efforts of the work packages, clinical departments, biomedical companies, and the Swedish patients’ organisation, *Mag*-och *tarmförbundet* [33].

**Figure 2.**
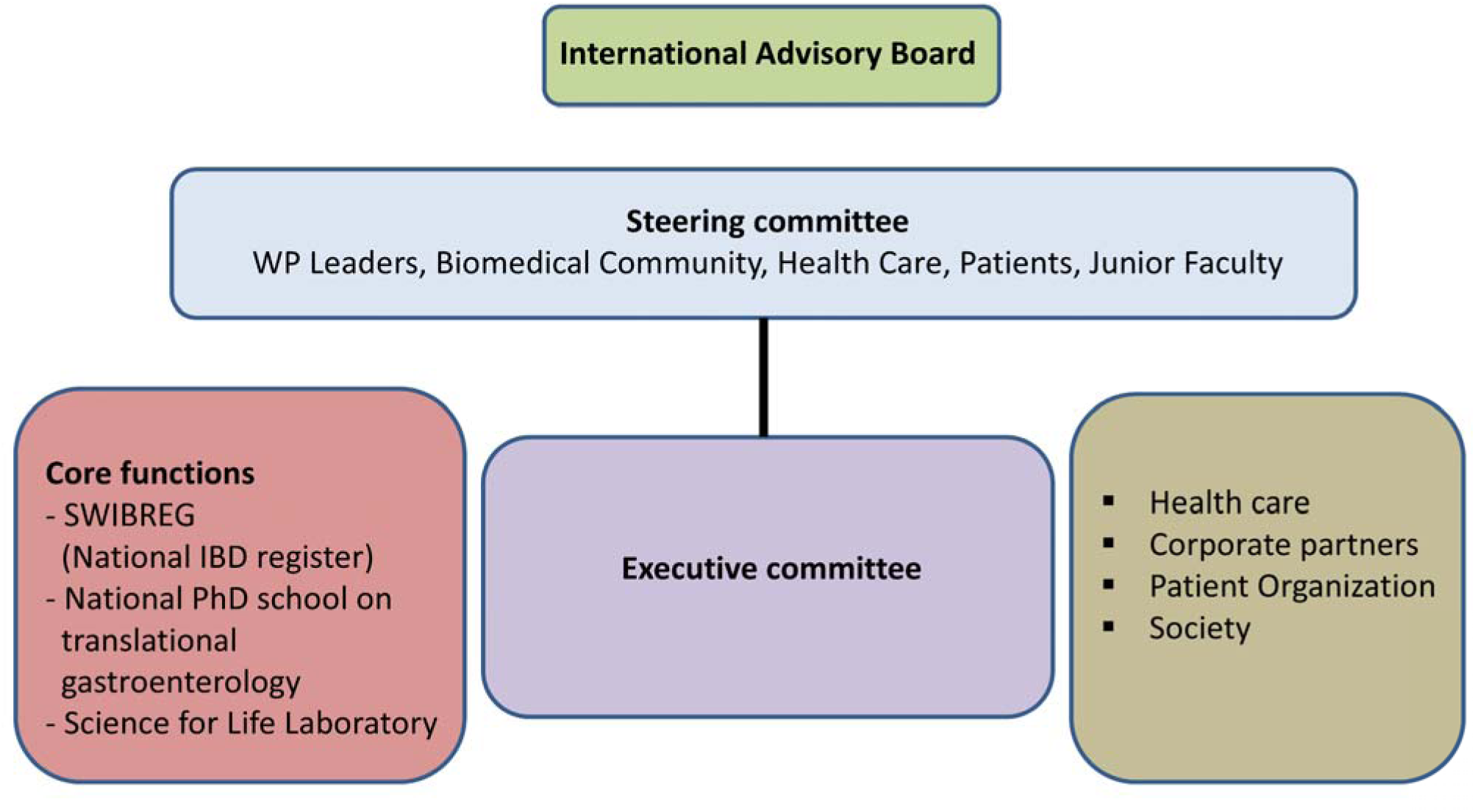
Managerial structure and the operational organisation of the multi-modal national study to identify biomarkers for diagnosis, therapy response and disease progression in inflammatory bowel disease (BIO IBD), where the Swedish inception cohort in IBD (SIC-IBD) represents an important resource.

The Steering Committee, responsible for the strategic leadership, is formed by representatives from involved universities, biomedical companies and the patient organisation, with equal numbers of basic scientists and clinicians, equal gender, and representation of young scientists. The Advisory Board, composed of internationally renowned representatives of academia and the biomedical industry, provides external review and input.

### Patient and Public Involvement statement

During the planning phase of the cohort study, patient representatives from the Swedish patient’s organisation, *Mag-och tarmförbundet* provided input on the study design, including the definition of the disease course during follow-up. Also, they assessed the burden of the collection of biological material and the time required for responding to the questionnaires in the study. Representatives of the patient organisation and the Swedish biomarker manufacturers in the executive committee also provided feedback during the recruitment process and follow-up of study participants. In addition to scientific reporting, key findings of the studies will be communicated to patient organisations, public health policymakers, and the public through various media and news activities. When disseminating the results, the recommendations of the International Committee of Medical Journal Editors (ICMJE)will be applied [34].

## FINDINGS TO DATE

At baseline, differences in smoking habits were observed between patients with Crohn’s disease, ulcerative colitis, and IBD-U. Active smoking was more prevalent in patients with Crohn’s disease, while a higher rate of former smokers was found in patients with ulcerative colitis. While hsCRP measurements were available for 90% of all patients and controls, only 63% of patients and controls had available FCP measures in the study. An additional 44 measurements of FCP at diagnosis were available from clinical routine, but these results were excluded from this analysis due to the use of different assays. Both FCP and hsCRP levels were higher in patients with Crohn’s disease and ulcerative colitis compared to symptomatic controls and healthy controls. However, while FCP levels were significantly elevated in IBD unclassified compared to symptomatic controls, the difference in hsCRP levels between these two groups was not statistically significant. No significant differences in FCP levels were observed between patients with Crohn’s disease and ulcerative colitis, but hsCRP levels were higher in Crohn’s disease patients. Significant differences in hsCRP and FCP levels were also found between symptomatic controls and healthy controls.

Preliminary results from analyses of serum proteins as well as findings from the analyses of faecal markers have highlighted the potential of these markers to differentiate between patients with IBD and symptomatic controls in this cohort, suggesting their diagnostic value [35,36]. Recent results from the analysis of anti-αvβ6 autoantibodies further suggested a high diagnostic value for ulcerative colitis of this marker [37]. Additionally, preliminary data indicate differences in mucosal proteins between patients developing an aggressive course of ulcerative colitis and those with an indolent course [38].

In line with previous studies [19,39], we employed a composite outcome to characterise the disease course within the first year after diagnosis, either as aggressive or indolent (Table 4). An aggressive course was defined by treatment refractoriness, the need for surgical resection, or in the case of Crohn’s disease, the development of severe complications such as strictures and fistulas. Based on this definition, 36% (52/143) of the patients with Crohn’s disease, 24% (49/201) of those with ulcerative colitis, and 4.2% (1/24) with IBD-U experienced an aggressive disease course within the first year after diagnosis (**Supplementary Table 1**). These results underscore that treatment failure rates and disease complications remain unacceptably high in IBD. Aggressive disease course was associated with more extensive ulcerative colitis, while no association was found between Crohn’s disease location and disease course.

## STRENGTHS AND LIMITATIONS

In this large nationwide multicentre inception cohort of newly diagnosed patients with IBD, a set of highly relevant biospecimens were collected at inclusion, and patients were prospectively followed. This cohort can be used to gain insights into key pathogenic mechanisms of IBD. Moreover, it provides a valuable resource for biomarker discoveries, particularly the identification of diagnostic biomarkers for IBD and prognostic markers for future disease courses.

This cohort is one of few large-scale IBD inception cohorts of adult patients with integrated biobanking [40–42], as most of the earlier initiatives have included paediatric patients only. Most previous attempts to identify diagnostic biomarkers in inception cohorts have used healthy controls or patients with non-inflammatory conditions (e.g., irritable bowel syndrome) as a control group. These settings do not reflect a diagnostic scenario where patients with gastrointestinal symptoms that are indicative of IBD are examined. Accordingly, evaluations based on these cohorts are likely to overestimate the diagnostic capacity of potential biomarkers [43]. However, this limitation is rarely acknowledged and may partly explain why promising biomarkers fail in replication attempts and do not make their way to the clinic. To allow the identification of diagnostic biomarkers, we have included patients with symptoms indicative of IBD but without any signs of the disease at diagnostic work-up or during follow-up (symptomatic controls) as a reference group. Thus, both cases with IBD and symptomatic controls represented an unselected sample of patients who were referred to secondary care for the suspicion of IBD. In addition, we included healthy controls as a second control group to gain insight into the aetiology of IBD and to characterise pathways related to disease pathogenesis.

In this cohort, we did not record the number of eligible patients who were not included, e.g. those who declined to or were not able to give informed consent or were not identified by study personnel. The absence of a population-based cohort design could be a limiting factor when interpreting associations between exposures and clinical outcomes within SIC-IBD. However, the proportion of IBD patients with Crohn’s disease in this study (39%), the European Crohn’s and Colitis Organization’s Epidemiological Committee (EpiCom) inception cohort (37%) and the recent population-based Inflammatory bowel disease in South-Eastern Norway cohort III (35%) were similar. Likewise, the proportions of ulcerative colitis (54%, 52% and 61%) and IBD-U (7%, 11% and 4%) were similar in the three cohorts. Suboptimal procedures in sample collection further limited the study. To enable inclusion at all centres, serum samples collected at centres with limited sample processing capacity were shipped overnight to a central biobank for centrifugation and biobanking. While measured faecal markers such as calprotectin are stable at room temperature for several days, microbiome analyses are sensitive to pre-analytic sample handling [44–46].

## COLLABORATION

We have established an inception cohort of newly diagnosed adult patients with IBD, symptomatic controls and healthy controls with integrated biobanking. Initiatives are underway to generate various multi–omics data from samples collected at baseline and follow-up visits. Analyses of single and multiple datasets, including integration of clinical variables, may discover novel diagnostic and prognostic biosignatures for IBD and its subtypes. Comparisons with healthy controls may also provide insights into IBD pathogenesis and progression mechanisms. We encourage collaborations with researchers from other cohorts and case-control studies to validate findings from the SIC-IBD cohort. Also, researchers can propose new collaborative initiatives and apply for access to data and biospecimens by submitting a proposal to the BIO IBD Executive Office. All proposals will be reviewed on scientific quality and methodology by the BIOIBD executive committee.

## Data Availability

Researchers can apply for access to data and biological specimens by submitting a proposal to the BIO IBD Executive Office, including a copy of their ethics approval. All proposals will be reviewed by the BIO IBD executive committee based on scientific quality and methodology. This procedure is installed to ensure that the clinical data and biological material are being requested for scientific purposes, in accordance with the informed consent signed by the participants, as the collected data and material cannot be used by purely commercial parties. Additionally, any transfer of data or biological material must comply with Swedish law and the European Union (EU) General Data Protection Regulation (GDPR).

## Acknowledgements

The authors would like to thank the SIC-IBD participants, involved nurses and physicians at the SIC-IBD study sites for their contribution. Also, the authors thank Åsa Smedberg and Anette Oskarsson for administrative support.

## Authors’ contributions

**Conception and design**: Jonas Halfvarson

**Drafting the manuscript**: Benita Salomon, Olle Grännö and Jonas Halfvarson

**Critical revision for intellectual content**: All authors

**Final approval**: All authors

**Accountable for all aspects of the work in ensuring that questions related to the accuracy or integrity of any part of the work are appropriately investigated and resolved**: All authors

## Funding

This work was supported by the Swedish Foundation for Strategic Research [RB13-0160 to J.H.], the Swedish Research Council [2020-02021 to J.H.], and the Örebro University Hospital research foundation [OLL-962042, OLL-974710, OLL-986849, OLL-1001470 to J.H.]. The funding sponsors had no role in the study’s design; collection, management, analysis, or interpretation of data; in writing of the protocol, or the decision to submit results for publication.

### Ethical approval

The SIC-IBD study was approved by the Regional Ethics Committees (Dnr 2010/313). Written informed consent was obtained from all participants before inclusion.

### Provenance and peer review

Not commissioned; externally peer-reviewed.

### Data sharing statement

Researchers can apply for access to data and biological specimens by submitting a proposal to the BIO IBD Executive Office, including a copy of their ethics approval. All proposals will be reviewed by the BIOIBD executive committee based on scientific quality and methodology. This procedure is installed to ensure that the clinical data and biological material are being requested for scientific purposes, in accordance with the informed consent signed by the participants, as the collected data and material cannot be used by purely commercial parties. Additionally, any transfer of data or biological material must comply with Swedish law and the European Union (EU) General Data Protection Regulation (GDPR).

---

**Supplementary Table 1.**

|  | Crohn's<br>disease<br>n=143 | ulcerative colitis n=201 | IBD-unclassified<br>n=24 |
| --- | --- | --- | --- |
| Aggressive disease, n (%) | 52 (36.4) | 45 (24.4) | 1 (4.2) |
| Indolent disease, n (%) | 79 (55.2) | 140 (69.7) | 22 (91.7) |
| Unknown*, n (%) | 12 (8.4) | 12 (6) | 1 (4.2) |

\*Missing information about clinical outcomes due to incomplete follow-up during the first year after diagnosis.

